# Dexmedetomidine as an Adjunctive Sedative in Patients Undergoing Endoscopic Submucosal Dissection: A Systematic Review and Meta-Analysis

**DOI:** 10.1101/2024.11.14.24317324

**Authors:** Hazem Abosheaishaa, Abdallfatah Abdallfatah, Abdelmalek Abdelghany, Arshia Sethi, Abdellatif Ismail, Doha Mohamed, Moataz Aboeldahb, Omar Abdelhalim, Islam Mohamed, Ahmed Y. Azzam, Muhammed Amir Essibayi, David J. Altschul, Mahmoud Nassar, Mohammad Bilal

## Abstract

**Introduction:** Endoscopic submucosal dissection (ESD) allows for curative en-bloc resection of dysplastic gastrointestinal (GI) tract lesions. However, it is associated with postoperative adverse events (AEs) such as pain, bleeding, and perforation. Dexmedetomidine, an α2-receptor agonist, has emerged as a promising adjunct sedative for ESD under moderate sedation, offering anxiolysis and analgesia. We conducted a systematic review and meta-analysis to evaluate its efficacy and safety for use in ESD.

**Methods:** A comprehensive systematic search was conducted across multiple databases, including Embase, Medline, Scopus, and Web of Science. Studies that involved ESD utilizing dexmedetomidine as an adjunctive medication in combination with other sedatives, were included. Data extraction and risk of bias assessment were independently performed by two reviewers. Meta-analysis was carried out with RevMan using a random-effects model.

**Results:** Eight studies were included in the final analysis. Dexmedetomidine showed no significant difference in en-bloc or complete resection rates compared to controls. Sedation and procedure times were similar between the two groups as well. Dexmedetomidine significantly reduced restlessness (OR 0.15, 95% CI:0.07 to 0.29) and increased bradycardia (OR 7.15, 95% CI 3.17 to 16.11) compared to controls. Upon subgroup analysis, Dexmedetomidine plus Propofol, and Dexmedetomidine plus Midazolam, revealed the same findings regarding restlessness and bradycardia compared to controls which confirmed the adjunctive effects of Dexmedetomidine.

**Conclusions:** Dexmedetomidine as an adjunctive sedative appears safe and effective in ESD, reducing restlessness without significant adverse events. The risk of bradycardia is increased, which may be reflective of reduced physiological stress. Future studies should explore optimal dosing and compare Dexmedetomidine with other sedatives in diverse populations.

## 1. Introduction

Endoscopic tumor resection is one of the most common modalities in GI tumor management. Endoscopic submucosal dissection (ESD) is considered superior to mucosal resection in view of offering complete resection with negative histological margins irrespective of the size of the original lesion [1] Despite these overwhelming advantages, ESD is associated with multiple postoperative complications including bleeding, postoperative perforation, and minor complications like abdominal pain, nausea, vomiting, and stricture which limits its use [2]. post-operative abdominal pain is a debilitating complication associated with ESD which is severely underestimated and results in decreased patient satisfaction and longer hospital stays. Studies show the incidence of postoperative pain in 44.9∼62.8% of patients, especially in the early post-operative period necessitating the use of aggressive pain management [3,4].

Dexmedetomidine is a new α2-receptor agonist that has anxiolytic, sedative, and analgesic properties which when used in combination with other anesthetics help lower their dose and also decrease postoperative opioid consumption and pain intensity [5,6]. A study done by Chang et al., also shows a better cardiovascular profile of dexmedetomidine as compared to propofol [7].

In our study, we reviewed the possible benefits of dexmedetomidine as an adjunct sedative perioperatively in patients undergoing ESD for GI adenomas and early-stage neoplastic lesions. We evaluated its efficacy by assessing variables like en-bloc resection, Complete resection, sedation time, procedure time, patient restlessness, and other adverse events.

## 2. Methods

### 2.1. Search Strategy and Data Extraction

A systematic search of relevant literature was conducted across multiple databases, including Embase, Scopus, Web of Science, Medline/PubMed, and Cochrane, from their inception to February 28, 2024. The search strategy utilized Boolean operators to combine terms related to the population, intervention, and outcomes of interest. The following search strategy was employed: (“endoscopic submucosal dissection” OR “ESD” OR “submucosal dissection”) AND (“dexmedetomidine” OR “dexmedetomidine” OR “sedative”) (**Appendix 1**). The search strategy aimed to identify studies investigating the use of dexmedetomidine as an adjunctive sedative in endoscopic submucosal dissection procedures. Our research adhered to the recommended guidelines for reporting systematic reviews and meta-analyses. The Preferred Reporting Items for Systematic Reviews and Meta-Analyses (PRISMA) checklist and Cochrane criteria were followed to ensure transparency and completeness in reporting [8,9].

Two independent reviewers screened titles, abstracts, and full-text articles for inclusion based on predefined eligibility criteria. Any disagreements were resolved through discussion or consultation with a third reviewer. Data extraction was conducted independently by two co-authors using a standardized data extraction form, with discrepancies resolved through consensus. Extracted data included study characteristics, patient demographics, details of the intervention and comparator, and outcomes of interest.

### 2.2. Inclusion Criteria and Study Outcomes

Studies eligible for inclusion in this meta-analysis were those focusing on patients who had gastrointestinal adenomas and early-stage neoplastic lesions eligible for endoscopic submucosal dissection (ESD) treatment. The intervention of interest was the use of dexmedetomidine as an adjunctive medication in combination with other sedatives in submucosal endoscopic dissection. There was no specific comparator for this review. The primary outcome of interest was the en-bloc resection. Secondary outcomes included Complete resection, sedative time, procedure time, restlessness, and different adverse events. Included study designs were randomized controlled trials (RCTs) and observational studies if applicable. Studies not written in English or with inadequate translation, Systematic reviews, Meta-analyses, Case reports, editorials, letters, or conference abstracts without full-text availability, animal studies, or studies conducted on non-human subjects were excluded.

### 2.3. Risk of Bias Assessment

The risk of bias and methodological quality of the included studies was assessed independently by two authors. The Cochrane risk-of-bias tool version 2, (ROB 2) was employed for RCTs. For observational studies, we used the Newcastle-Ottawa Scale, any discrepancies were resolved through discussion or consultation with a third reviewer [10].

### 2.4. Statistical Analysis

A meta-analysis was conducted using Review Manager 5.4 (Cochrane Collaboration, Copenhagen, The Nordic Cochrane Centre). Given the anticipated heterogeneity in study designs and populations, a random-effects model was utilized. Summary measures were expressed as pooled odds ratios (OR) with corresponding 95% confidence intervals (CI) for proportional variables and mean differences with corresponding 95% CIs for continuous variables. Statistical significance was set at a p-value <0.05. Heterogeneity was assessed using the I2 statistic, with an I2 value of ≥50% indicating significant heterogeneity defined by the Cochrane Handbook for systematic reviews [11].

## 3. Results

### 3.1. Search results

The initial search retrieved 216 studies, 104 of them underwent title and abstract screening, and 25 full texts were assessed for inclusion. Eight studies were included in our final analysis (**Figure 1**).

**Figure 1:**
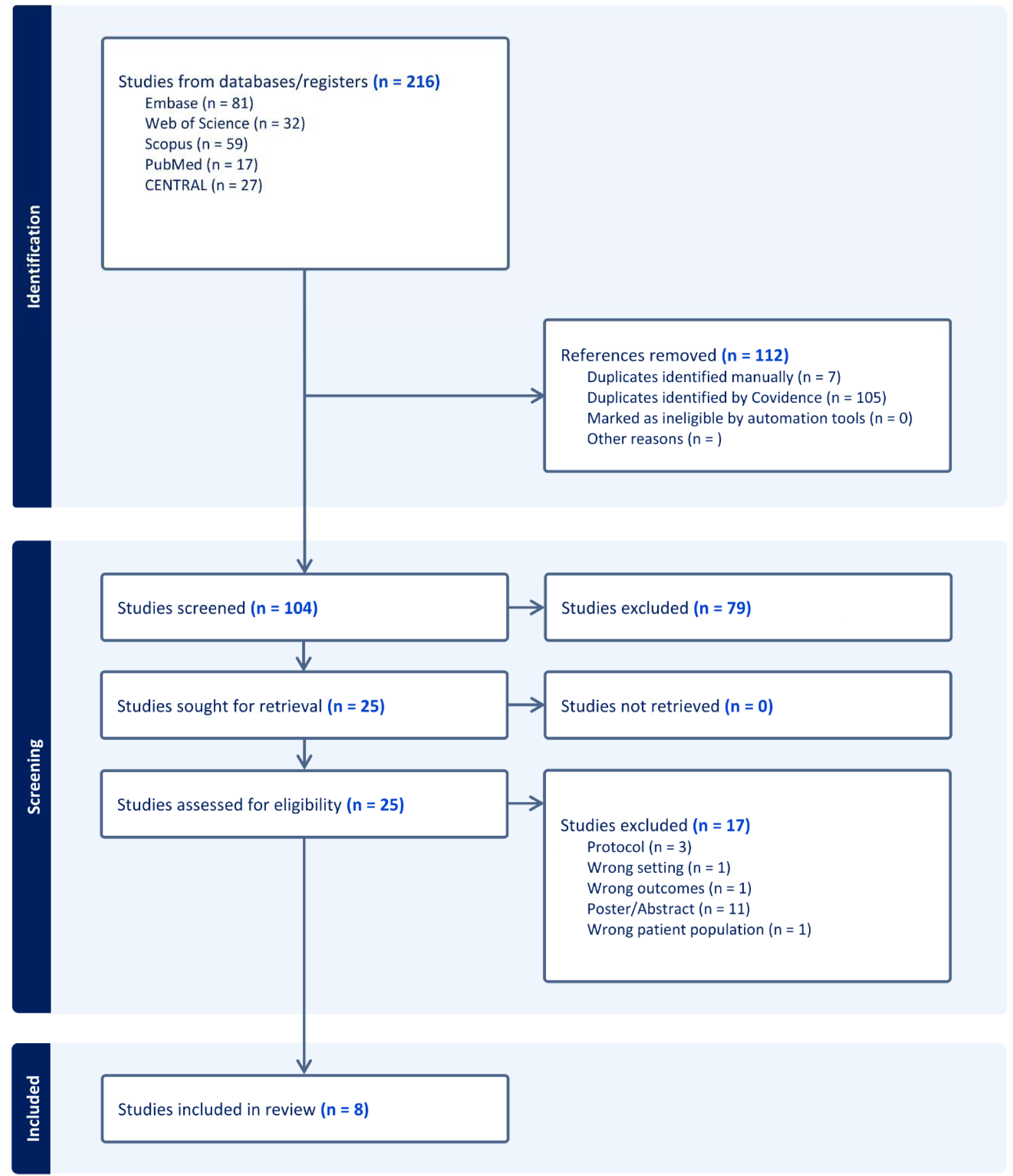
PRISMA flow diagram of the search strategy.

### 3.2. Study and patient characteristics

A total of 836 patients were included in our meta-analysis. Of the 836 patients, 412 (49.2%) were assigned to the Dexmedetomidine group, whereas 424 (50.7%) were assigned to the placebo group.

The included eight studies’ characteristics are displayed in (**Table 1**).

**Table 1:**
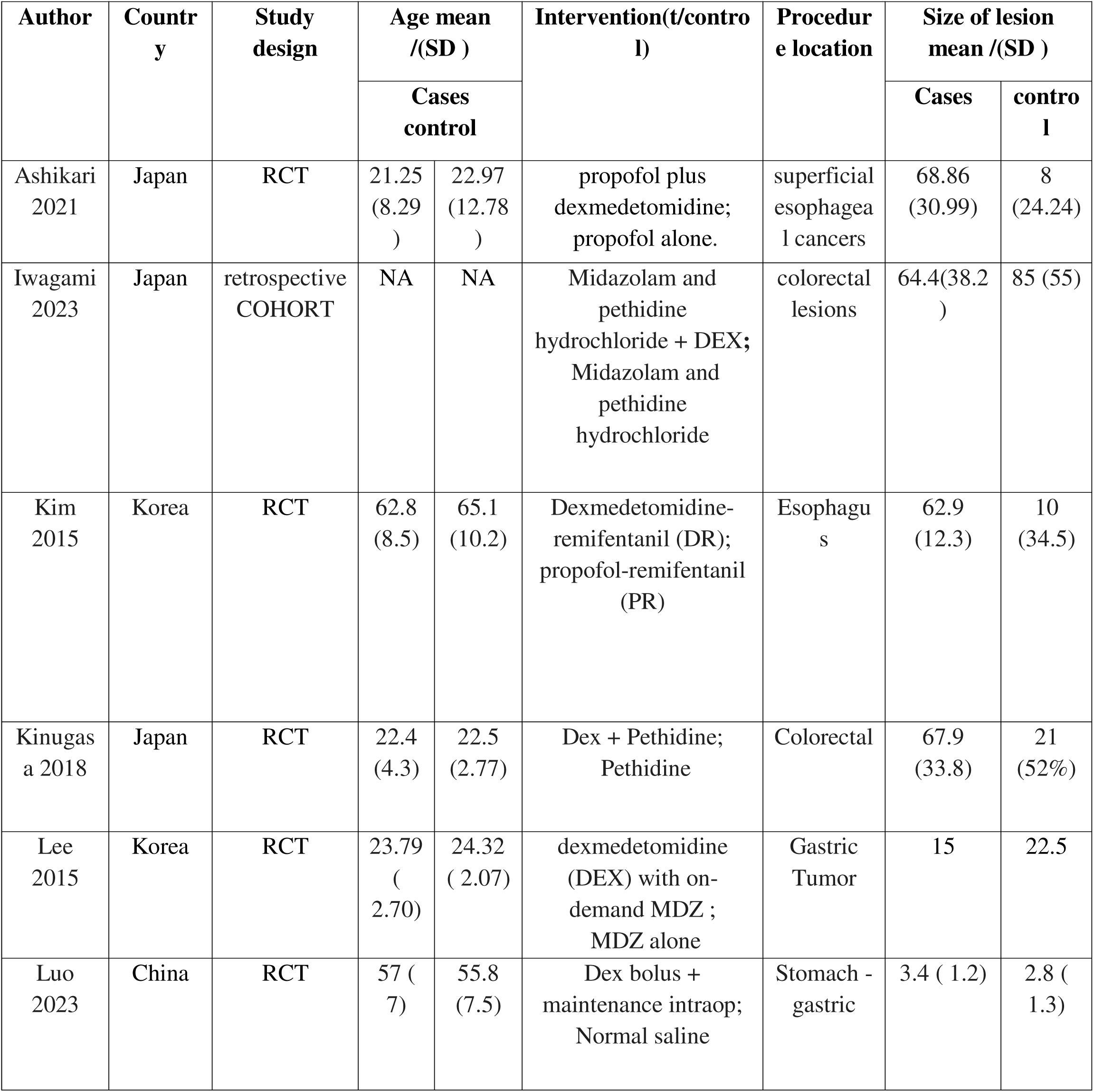
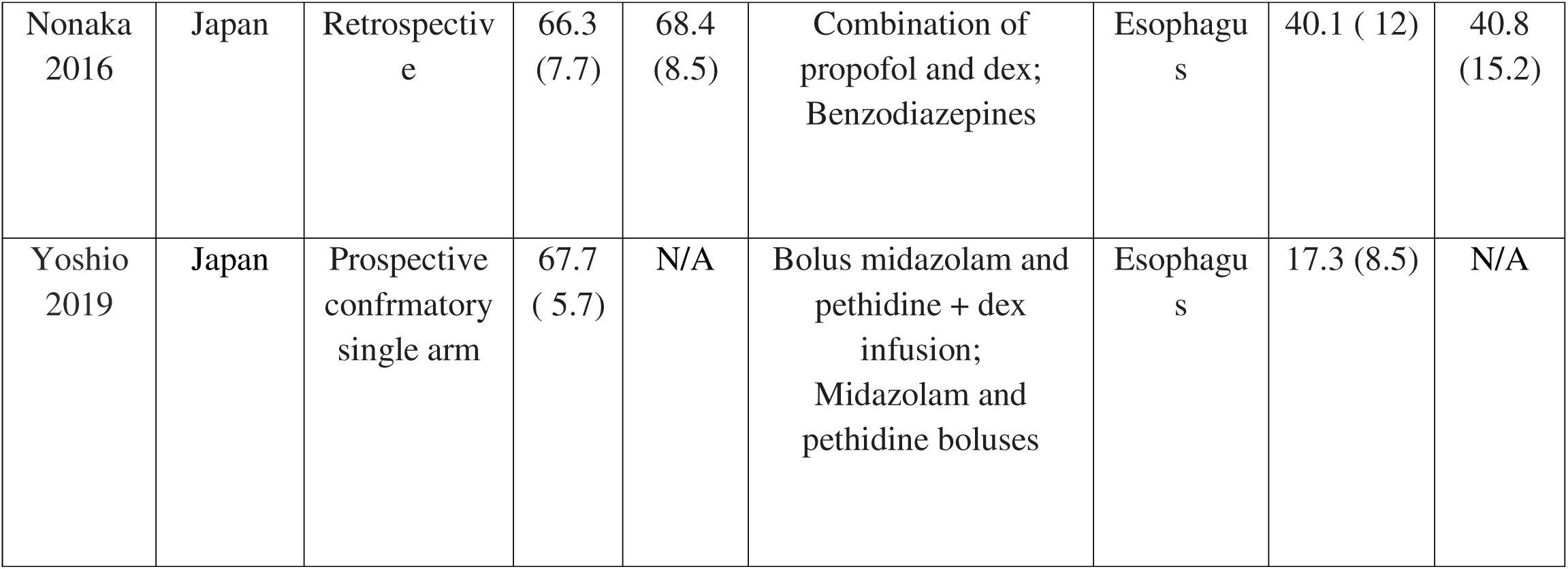
Baseline Characteristics of The Included Studies.

**Tables Legend:**
| Author | Country | Study design | Age mean /(SD ) |  | Intervention(t/control) | Procedure location | Size of lesion mean /(SD ) |  |
| --- | --- | --- | --- | --- | --- | --- | --- | --- |
|  |  |  | Cases control |  |  |  | Cases | control |
| Ashikari 2021 | Japan | RCT | 21.25 (8.29 ) | 22.97 (12.78 ) | propofol plus dexmedetomidine; propofol alone. | superficial esophageal cancers | 68.86 (30.99) | 8 (24.24) |
| Iwagami 2023 | Japan | retrospective COHORT | NA | NA | Midazolam and pethidine hydrochloride + DEX; Midazolam and pethidine hydrochloride | colorectal lesions | 64.4(38.2 ) | 85 (55) |
| Kim 2015 | Korea | RCT | 62.8 (8.5) | 65.1 (10.2) | Dexmedetomidine-remifentanil (DR); propofol-remifentanil (PR) | Esophagus | 62.9 (12.3) | 10 (34.5) |
| Kinugasa 2018 | Japan | RCT | 22.4 (4.3) | 22.5 (2.77) | Dex + Pethidine; Pethidine | Colorectal | 67.9 (33.8) | 21 (52%) |
| Lee 2015 | Korea | RCT | 23.79 ( 2.70) | 24.32 ( 2.07) | dexmedetomidine (DEX) with on-demand MDZ ; MDZ alone | Gastric Tumor | 15 | 22.5 |
| Luo 2023 | China | RCT | 57 ( 7) | 55.8 (7.5) | Dex bolus + maintenance intraop; Normal saline | Stomach - gastric | 3.4 ( 1.2) | 2.8 ( 1.3) |
| Nonaka<br>2016 | Japan | Retrospective | 66.3<br>(7.7) | 68.4<br>(8.5) | Combination of propofol and dex; Benzodiazepines | Esophagus | 40.1 ( 12) | 40.8<br>(15.2) |
| Yoshio<br>2019 | Japan | Prospective<br>confirmatory<br>single arm | 67.7<br>( 5.7) | N/A | Bolus midazolam and pethidine + dex infusion;<br>Midazolam and pethidine boluses | Esophagus | 17.3 (8.5) | N/A |

### 3.3. Quality of included studies

Quality assessment of included studies was assessed using (the Cochrane RoB 2 tool) for Randomized clinical trials. Four studies had a total low risk of bias, and one study had a moderate risk of bias. Another three Cohort studies were assessed using the Newcastle–Ottawa Scale with a low risk of bias (**Appendix 2**).

### 3.4. Meta-analysis outcomes

#### 1 En-bloc resection

The data from 7 studies were analyzed for En-bloc resection, the odds ratio was 1.45 with a 95% confidence interval (CI) of 0.47 to 4.41 which revealed no significant difference between the two groups (p=0.52) at random effect as shown in **Figure 2**.

**Figure 2:**
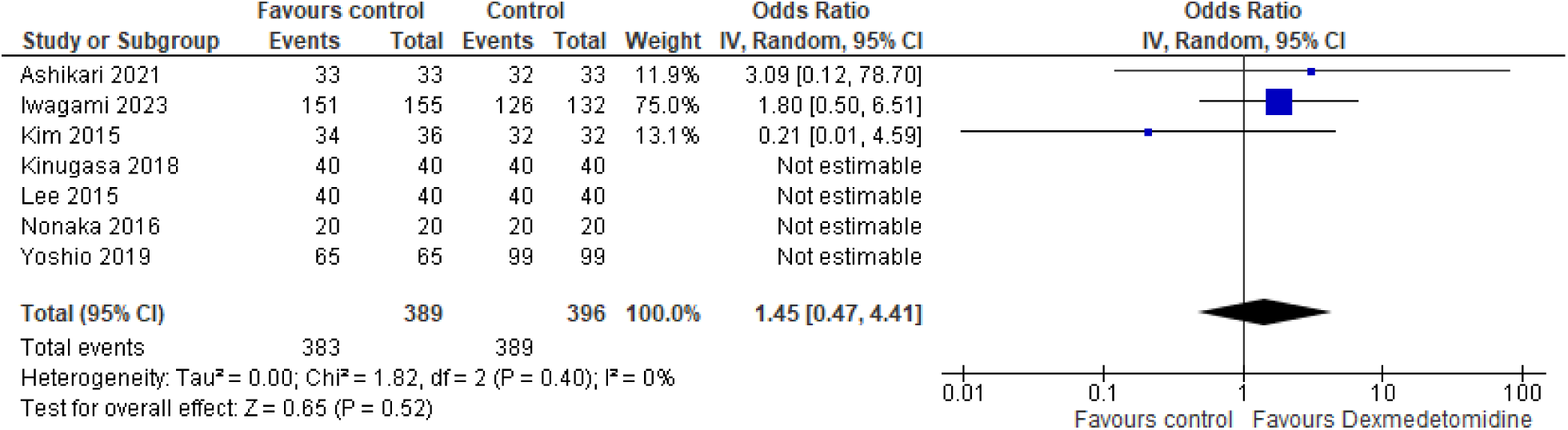
En-bloc resection.

#### 2 Complete resection

Three studies reported a complete resection rate, and the odds ratio was 0.62 with a 95% confidence interval (CI) of 0.21 to 1.80 which revealed no significant difference between the two groups (p=0.38) as shown in **Figure 3**.

**Figure 3:**
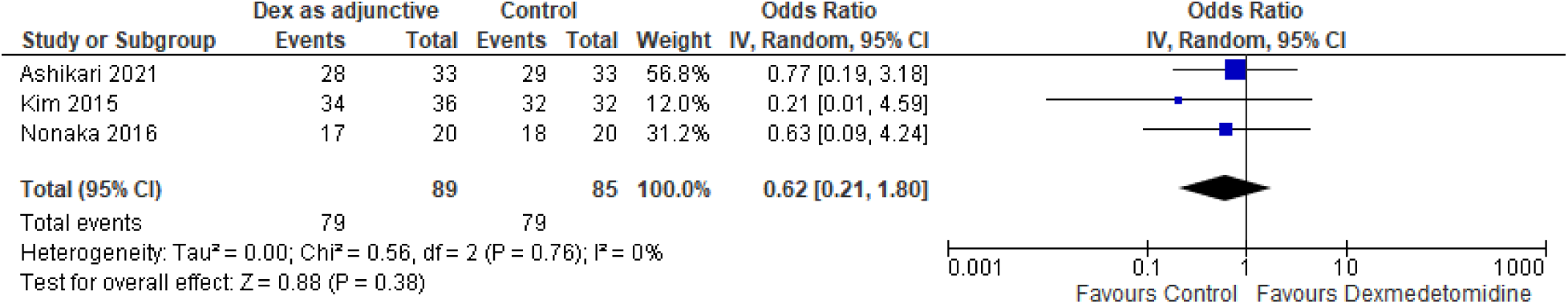
Complete resection.

#### 3 Sedation time

The pooled results from four studies reporting on sedation time revealed that there was no significant difference between the two groups, as shown in **Figure 4** (MD: 6.81, 95% CI: −2.25-15.87; I^2^ 0%; P=0.14).

**Figure 4:**
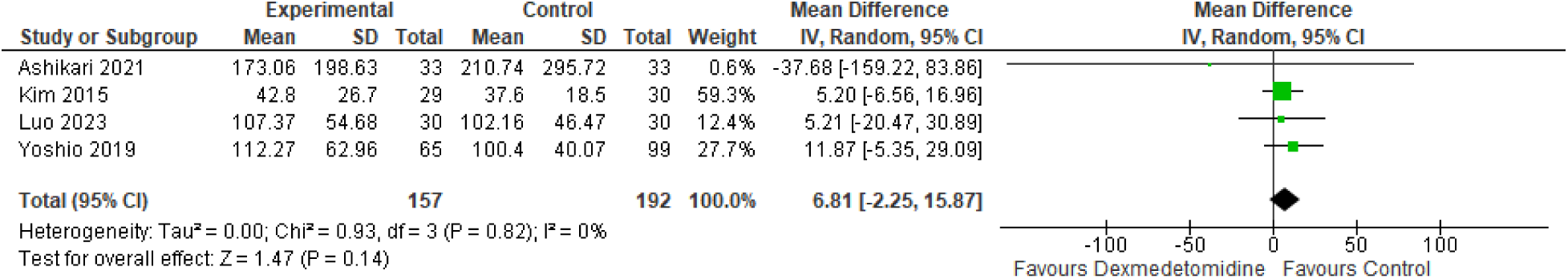
Sedation time.

#### 4 Procedure Time

Five studies reporting on procedure time revealed that there was no significant difference between the two groups as shown in **Figure 5** (MD: 3.21, 95% CI: −6.32-12.74; I^2^ 0%; P=0.51).

**Figure 5:**
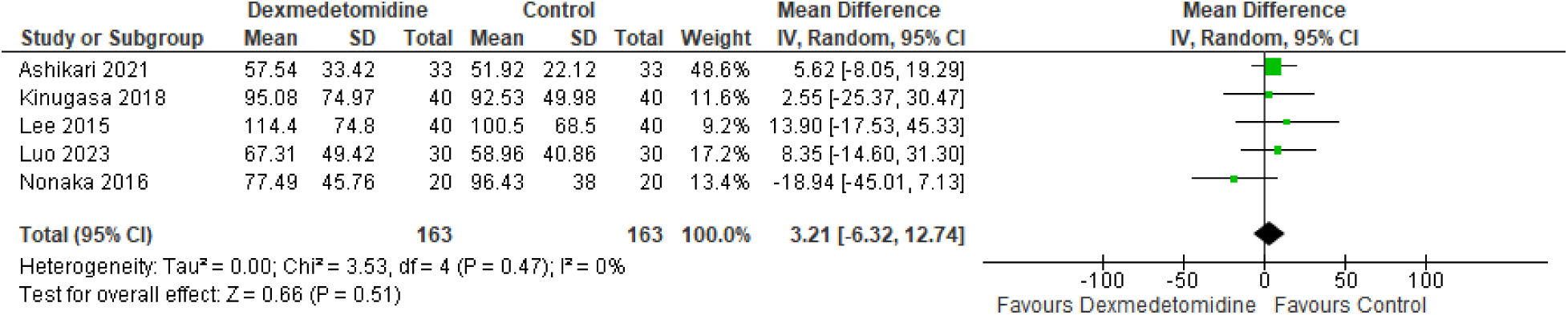
Procedure Time.

#### 5 Restlessness

Four studies reported a restlessness rate, the odds ratio was 0.15 with a 95% confidence interval (CI) of 0.07 to 0.29 which revealed a significant difference between the two groups (p<0.00001). as shown in **Figure 6**.

**Figure 6:**
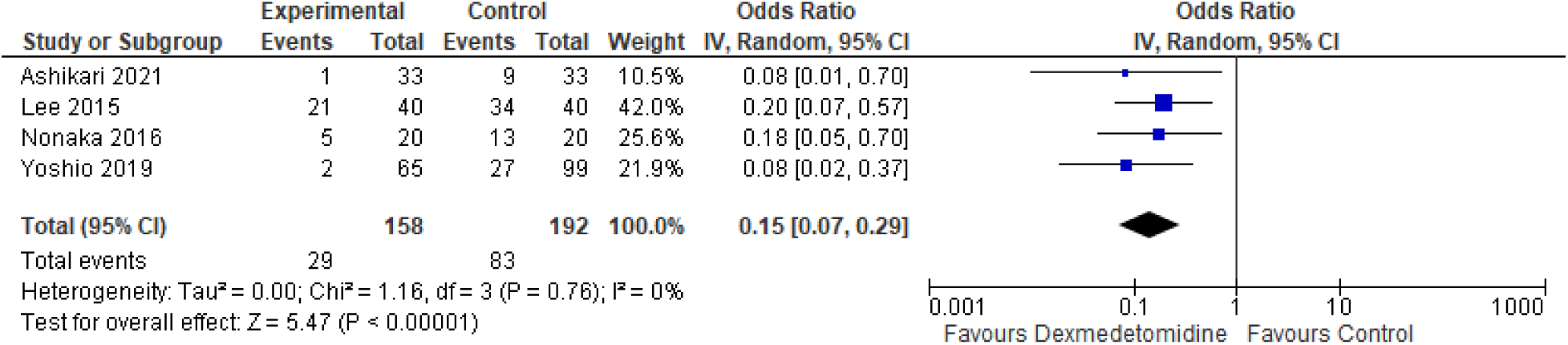
Restlessness.

#### 6 Bradycardia

Seven studies reported the bradycardia rate, and the odds ratio was 7.15 with a 95% confidence interval (CI) of 3.17 to 16.11 which revealed a significant difference between the two groups (p<0.00001) as shown in **Figure 7**.

**Figure 7:**
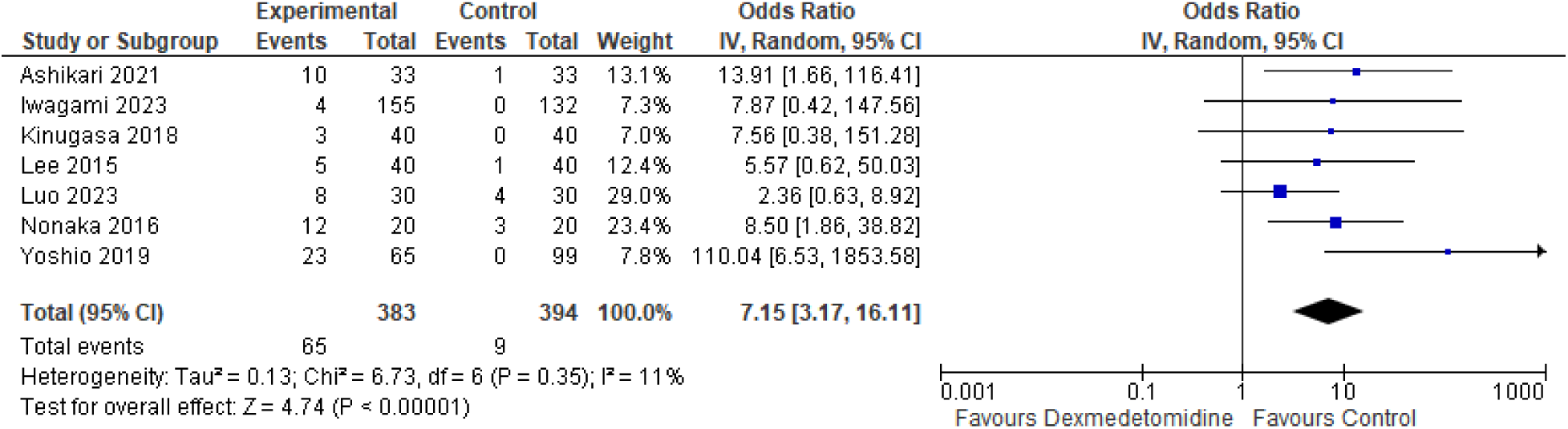
Bradycardia.

#### 7 Hypoxia

Four studies reported the Hypoxia rate and the odds ratio was 0.95 with a 95% confidence interval (CI) of 0.38 to 2.36 which revealed no significant difference between the two groups (p=0.91) as shown in **Figure 8**.

**Figure 8:**
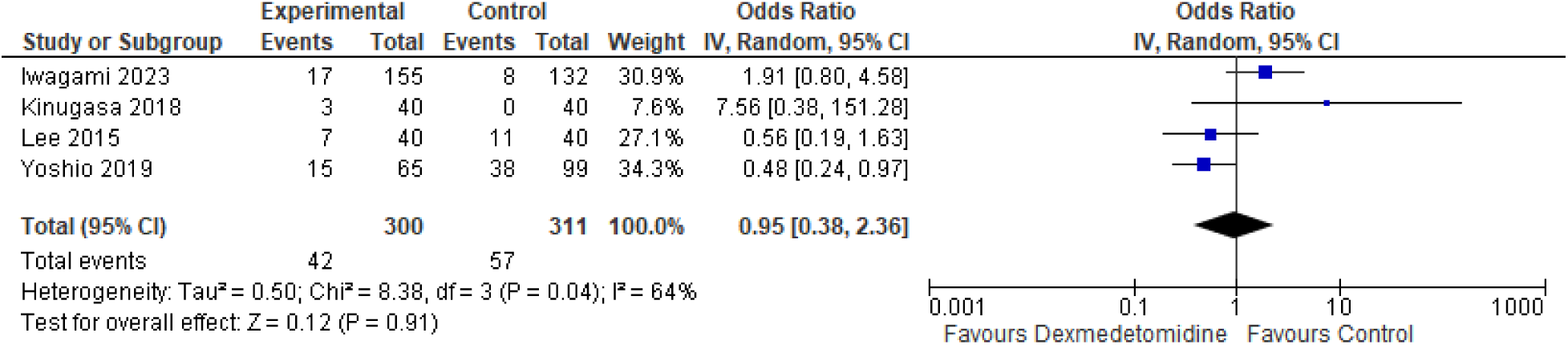
Hypoxia.

#### 8 Hypotension

Seven studies reported the Hypotension rate and the odds ratio was 2.73 with a 95% confidence interval (CI) of 0.79 to 9.43 which revealed no significant difference between the two groups (p=0.11) as shown in **Figure 9**.

**Figure 9:**
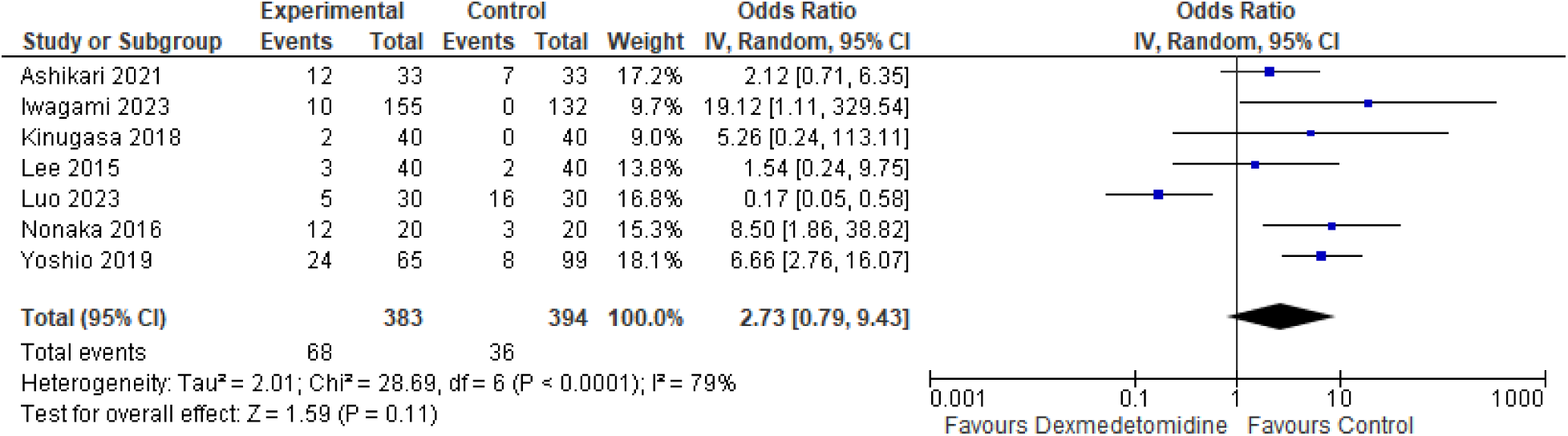
Hypotension.

#### 9 Perforation

Five studies reported the perforation rate and the odds ratio was 0.51 with a 95% confidence interval (CI) of 0.05 to 5.44 which revealed no significant difference between the two groups (p=0.58) as shown in **Figure 10**.

**Figure 10:**
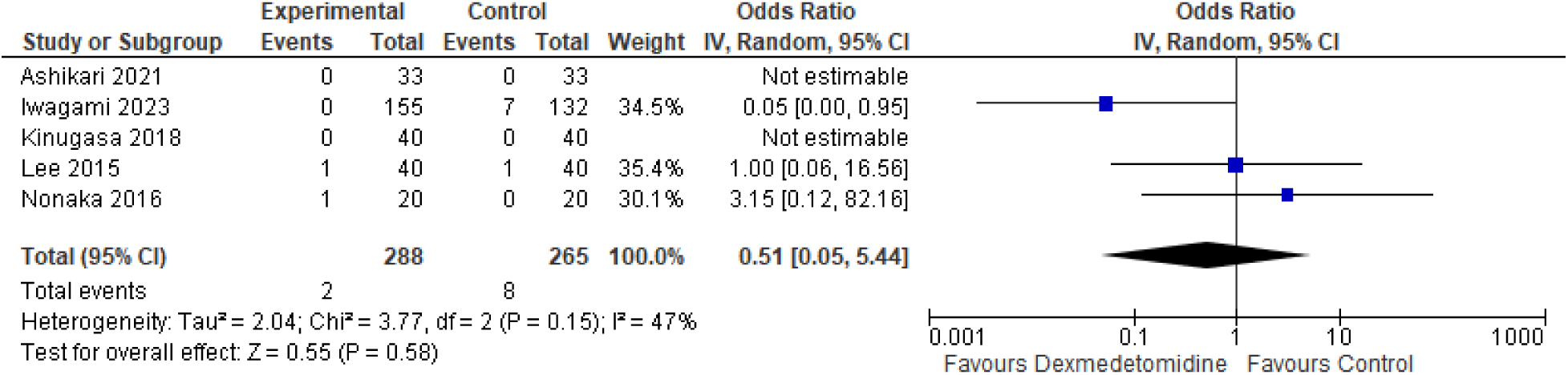
Perforation.

#### 10 Bleeding

Three studies reported the Bleeding rate, the odds ratio was 0.41 with a 95% confidence interval (CI) of 0.12 to 1.39 which revealed no significant difference between the two groups (p=0.15) .as shown in **Figure 11**.

**Figure 11:**
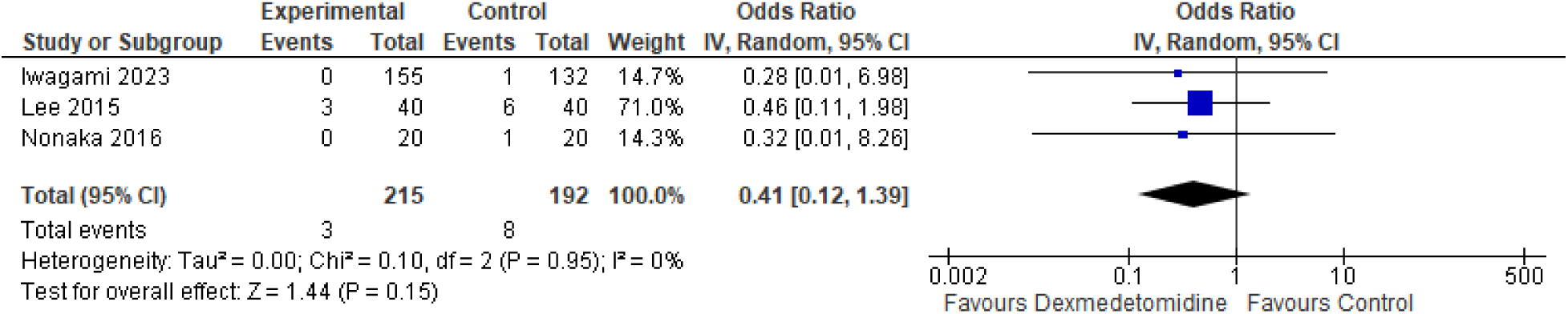
Bleeding.

### 3.5. Subgroup Analysis Outcomes

#### 1 Dexmedetomidine + Propofol

##### 1.1. En-bloc resection (Dexmedetomidine + Propofol)

Two studies reported en-bloc resection rate for Dexmedetomidine in combination with propofol, the odds ratio was 3.09 with a 95% confidence interval (CI) of 0.12 to 78.70 which revealed no significant difference between the two groups (p=0.49) as shown in **Figure 12**.

**Figure 12:**
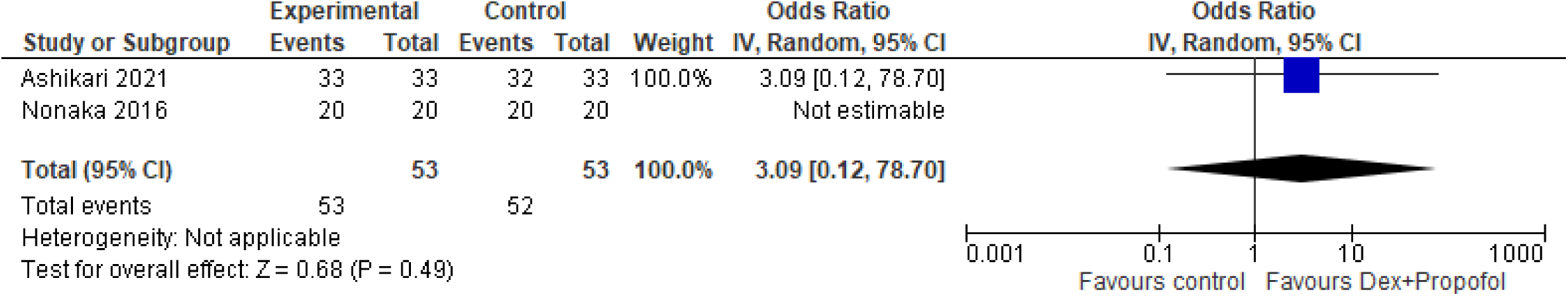
En-bloc resection (Dexmedetomidine + Propofol).

##### 1.2. Complete resection (Dexmedetomidine + Propofol)

Two studies reported a complete resection rate for Dexmedetomidine in combination with propofol, the odds ratio was 0.72 with a 95% confidence interval (CI) of .23 to 2,24 which revealed no significant difference between the two groups (p=0.57) as shown in **Figure 13**.

**Figure 13:**
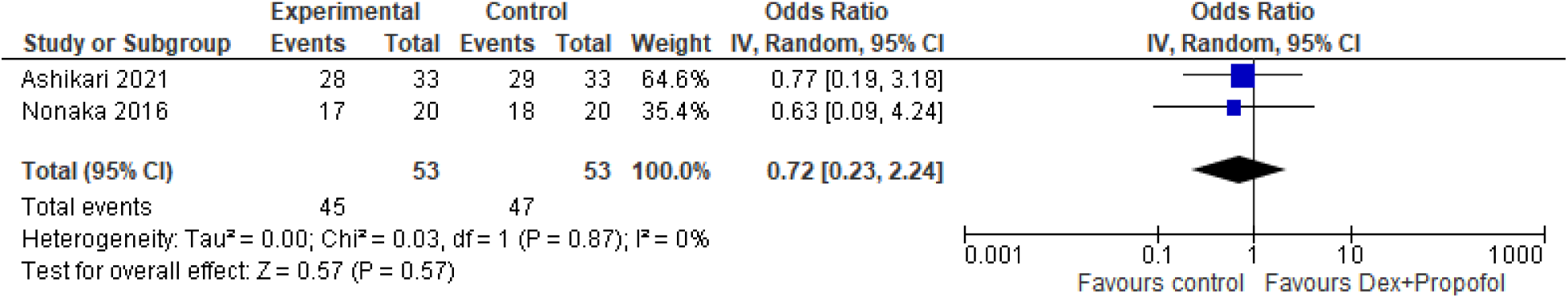
Complete resection (Dexmedetomidine + Propofol).

##### 1.3. Procedure Time (Dexmedetomidine + Propofol)

Two studies reporting on procedure time for Dexmedetomidine in combination with propofol revealed that there was no significant difference between the two groups as shown in **Figure 14** (MD: −4.05, 95% CI: −27.57-19.47; I^2^=63%; P=0.74).

**Figure 14:**
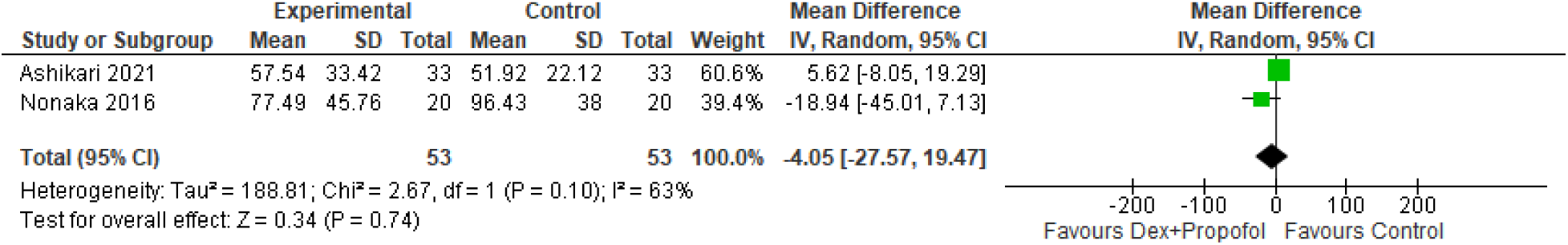
Procedure Time (Dexmedetomidine + Propofol).

##### 1.4. Restlessness (Dexmedetomidine + Propofol)

Two studies reported Restlessness for Dexmedetomidine in combination with propofol, the odds ratio was 0.14 with a 95% confidence interval (CI) of 0.05 to 0.45 which revealed a significant difference between the two groups (p=0.0009) as shown in **Figure 15**.

**Figure 15:**
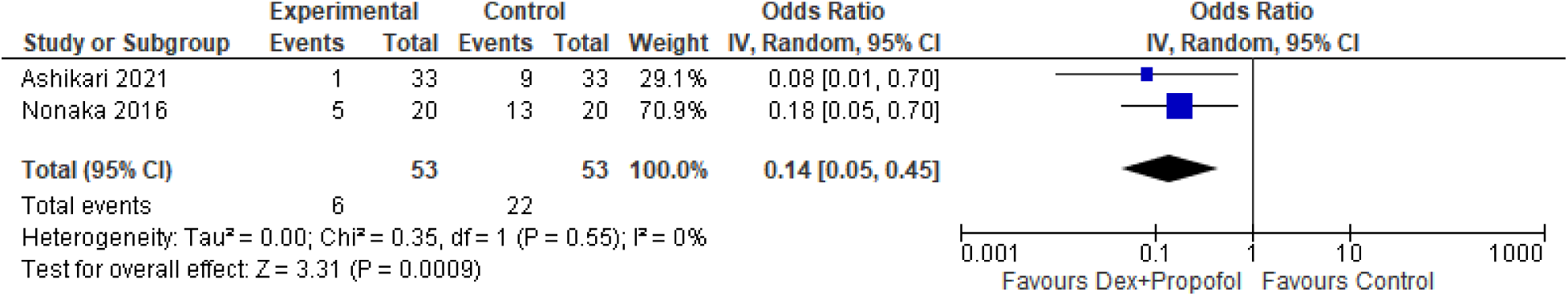
Restlessness (Dexmedetomidine + Propofol).

##### 1.5. Bradycardia (Dexmedetomidine + Propofol)

Two studies reported Bradycardia for Dexmedetomidine in combination with propofol, the odds ratio was 10.04 with a 95% confidence interval (CI) of 2.92 to 34.54 which revealed a significant difference between the two groups (p=0.0003) as shown in **Figure 16**.

**Figure 16:**
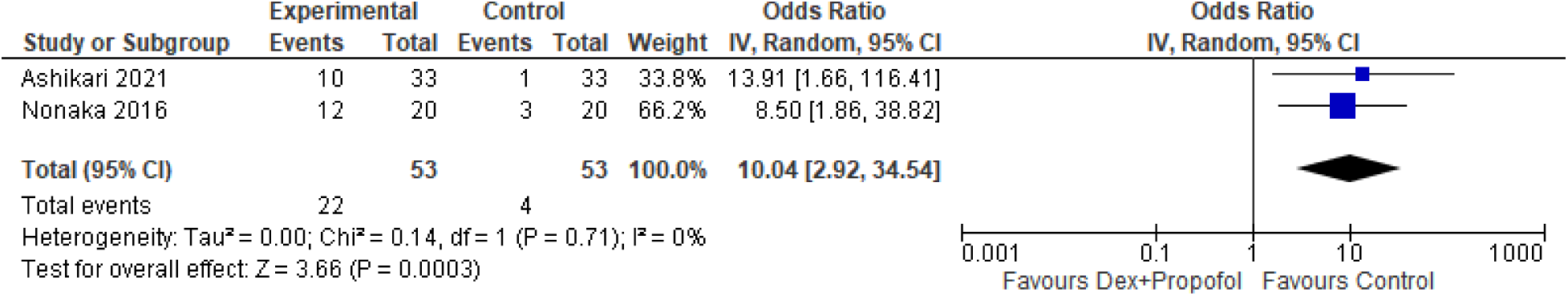
Bradycardia (Dexmedetomidine + Propofol).

##### 1.6. Hypoxemia (Dexmedetomidine + Propofol)

Two studies reported hypoxemia for Dexmedetomidine in combination with propofol, the odds ratio was 0.28 with a 95% confidence interval (CI) of 0.11 to 0.71 which revealed a significant difference between the two groups (p=0.007) as shown in **Figure 17**.

**Figure 17:**
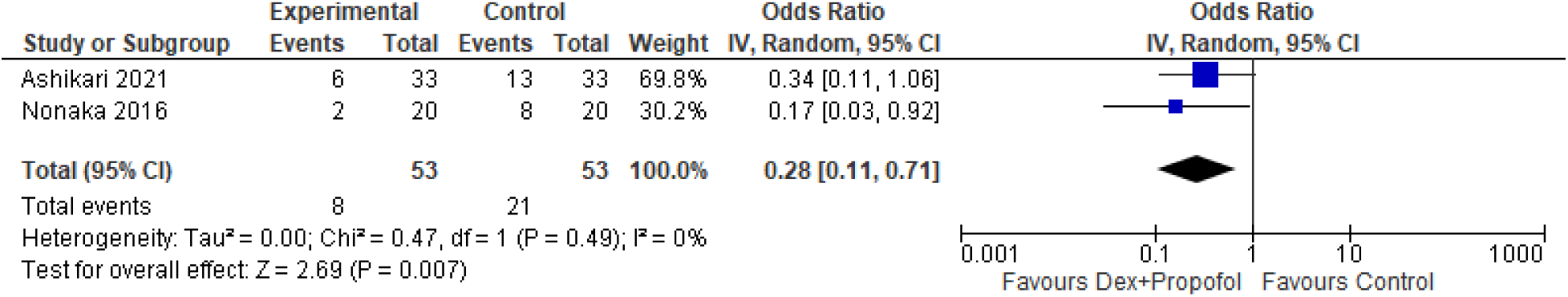
Hypoxemia (Dexmedetomidine + Propofol).

##### 1.7. Hypotension (Dexmedetomidine + Propofol)

Two studies reported Hypotension for Dexmedetomidine in combination with propofol, the odds ratio was 3.83 with a 95% confidence interval (CI) of 1.00 to 14.69 which revealed a significant difference between the two groups (p=0.05) as shown in **Figure 18**.

**Figure 18:**
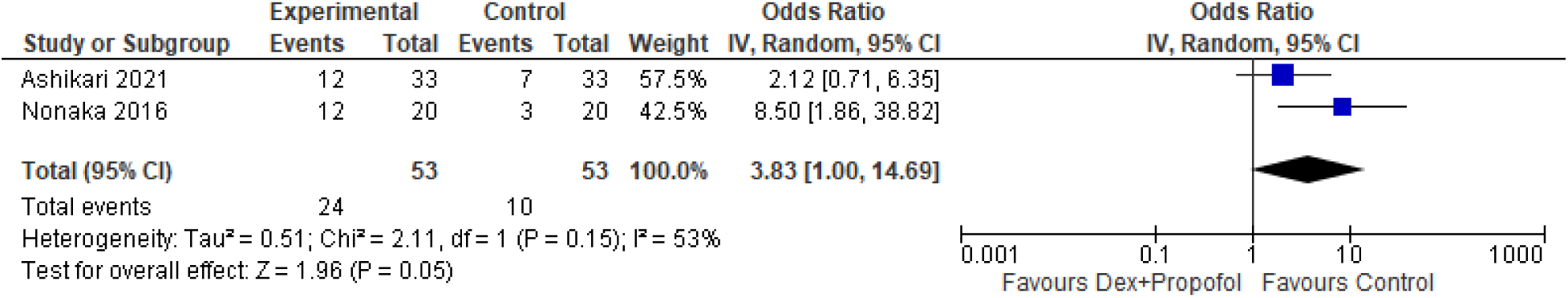
Hypotension (Dexmedetomidine + Propofol).

#### 2 Dex + Midazolam

##### 2.1. En-bloc resection (Dex + Midazolam)

Two studies reported en-bloc resection for Dexmedetomidine in combination with Midazolam, the odds ratio was 1.80 with a 95% confidence interval (CI) of 0.50 to 6.51 which revealed no significant difference between the two groups (p=0.37) .as shown in **Figure 19**.

**Figure 19:**
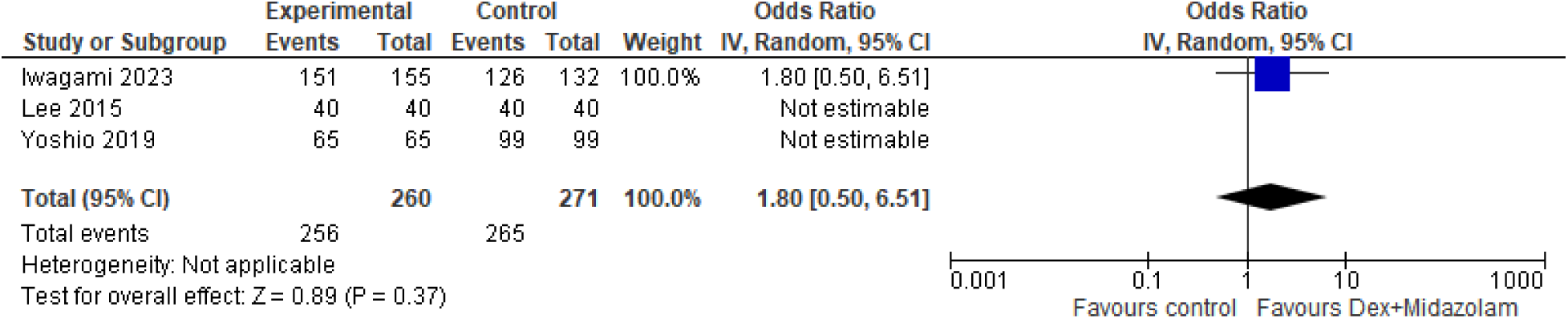
En-bloc resection (Dex + Midazolam).

##### 2.2. Restlessness (Dex + Midazolam)

Two studies reported Restlessness for Dexmedetomidine in combination with Midazolam, the odds ratio was 0.15 with a 95% confidence interval (CI) of 0.06 to 0.35 which revealed a significant difference between the two groups (p < 0.0001) as shown in **Figure 20**.

**Figure 20:**
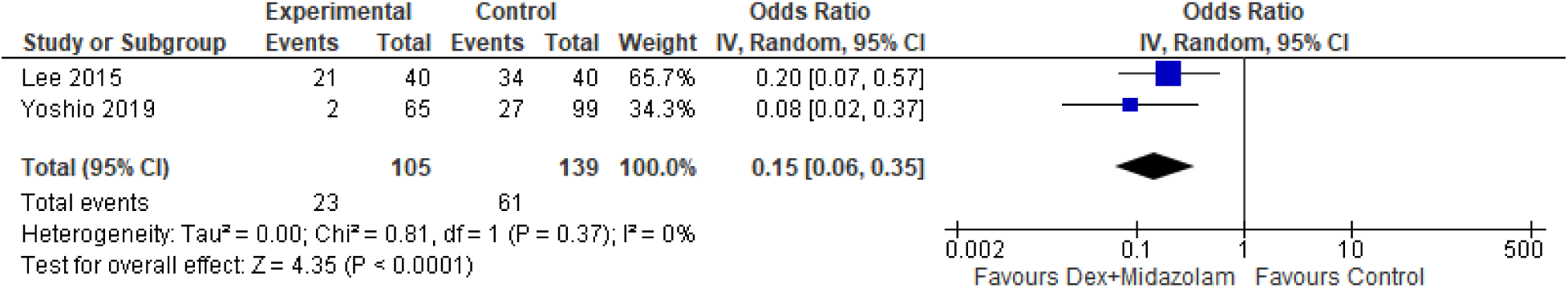
Restlessness (Dex + Midazolam).

##### 2.3. Bradycardia (Dex + Midazolam)

Two studies reported Bradycardia for Dexmedetomidine in combination with Midazolam, the odds ratio was 14.97 with a 95% confidence interval (CI) of 2.44 to 91.68 which revealed a significant difference between the two groups (p=0.003) as shown in **Figure 21**.

**Figure 21:**
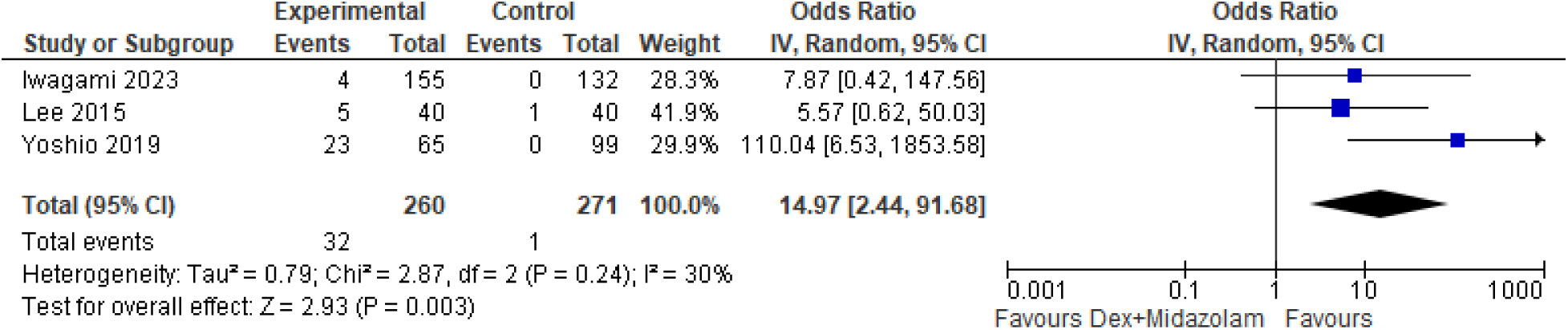
Bradycardia (Dex + Midazolam).

##### 2.4. Hypoxia (Dex + Midazolam)

Two studies reported Hypoxia for Dexmedetomidine in combination with Midazolam, the odds ratio was 0.80 with a 95% confidence interval (CI) of 0.33 to 1.94 which revealed no significant difference between the two groups (p=0.62) as shown in **Figure 22**.

**Figure 22:**
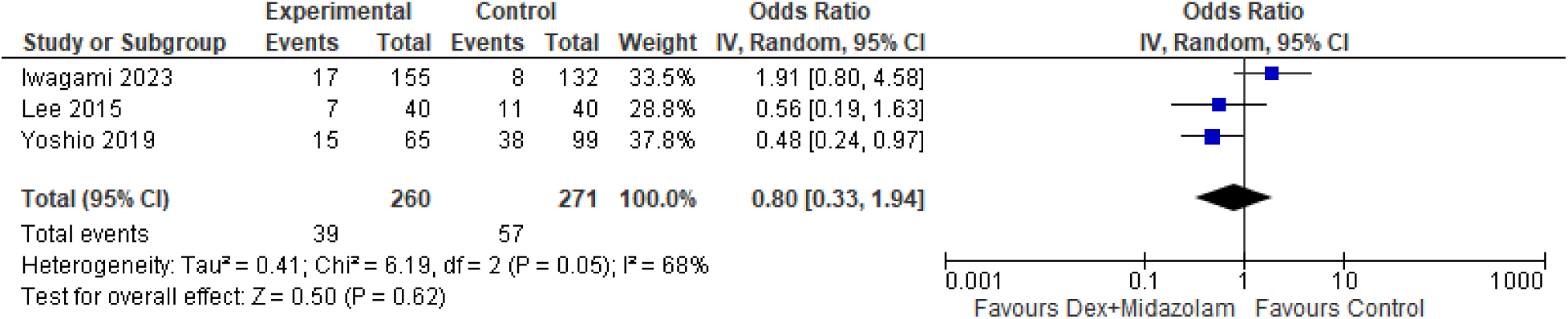
Hypoxia (Dex + Midazolam).

##### 2.5. Bleeding (Dex + Midazolam)

Two studies reported Bleeding for Dexmedetomidine in combination with Midazolam, the odds ratio was 0.42 with a 95% confidence interval (CI) of 0.11 to 1.60 which revealed no significant difference between the two groups (p=0.20) as shown in **Figure 23**.

**Figure 23:**
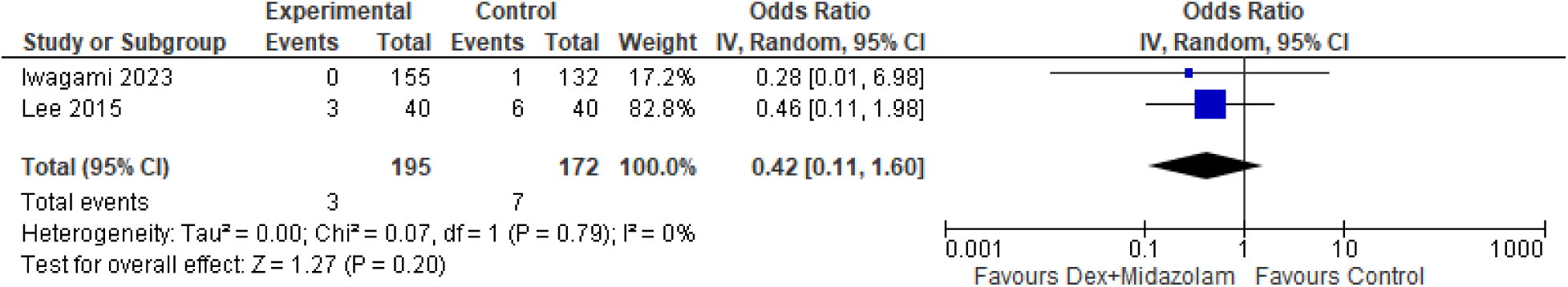
Bleeding (Dex + Midazolam).

##### 2.6. Perforation (Dex + Midazolam)

Two studies reported Perforation for Dexmedetomidine in combination with Midazolam, the odds ratio was 0.24 with a 95% confidence interval (CI) of 0.01 to 4.13 which revealed no significant difference between the two groups (p=0.32) as shown in **Figure 24**.

**Figure 24:**
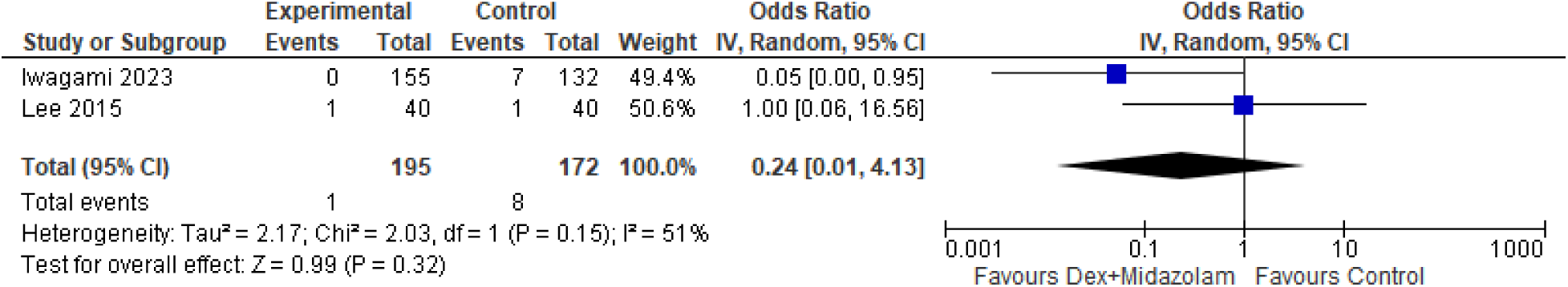
Perforation (Dex + Midazolam).

## 4. Discussion

Although the debilitating pain associated with ESD warrants aggressive pain management, physicians are hesitant due to the possibility of masking the pain of perforation. This not only causes patient discomfort but also increases the burden on healthcare by prolonging discharge time [2,4]. A study done by Seiichiro et al. shows an increased incidence of metachronous gastric cancer in patients who underwent curative ESD of early gastric cancer [12]. These warrant further endoscopic surveillance and possible repeat ESD. However, poorly managed post-operative pain increases apprehension in patients for further endoscopic procedures. As previously described, a few studies have been done describing the incidence of postoperative pain after ESD but there is no consensus on the management of the said pain. Studies done by Lee and Kim recommend a single dose of dexamethasone or postoperative local bupivacaine and triamcinolone [13,14].

In our study, we found a significant reduction in restlessness and bradycardia associated with dexmedetomidine highlighting its potential as an effective sedative agent for endoscopic procedures. We also observed a statistically significant decrease in tachycardia which could indicate less anxiety and pain thus providing a more comfortable sedative experience for the patients. These properties could be attributed to its selective alpha 2 adrenergic agonist and sympatholytic properties [15]. The non-significant difference found in en bloc resection rates between dexmedetomidine, and the comparator groups alleviated concerns regarding the influence of the sedation regimen on the technical aspects of the procedure [16].

The subgroup analysis also revealed better outcomes particularly in terms of reduced restlessness and bradycardia, with Dexmedetomidine in combination with Midazolam compared to other combinations. The anxiolytic and amnestic properties of Midazolam, coupled with the sedative and analgesic effects of Dexmedetomidine, may offer superior patient comfort and procedural tolerance. Additionally, considering the favorable safety profile of Midazolam in terms of respiratory depression compared to Propofol, this combination presents a compelling option for optimizing sedation strategies in endoscopic settings [17].

Our review suggests that dexmedetomidine is an effective sedative agent for ESD. Lee et al. [18] on the other hand, compared the outcomes of sedation using dexmedetomidine infusion plus on-demand midazolam versus sedation using midazolam infusion plus on-demand midazolam. They concluded that the sedation effect of dexmedetomidine with midazolam was superior to the sedation effect with midazolam alone. Furthermore, four studies reported sedation time as an outcome, while five studies reported procedure time as an outcome, **Figure 4** and **Figure 5**. The pooled results from these studies showed no statistically significant difference in sedation or procedure time between the dexmedetomidine and the control group. Despite the significant reduction in intraoperative restlessness in the dexmedetomidine group in our review, as mentioned above, this did not translate into a shorter sedation or procedure time, (**Figure 6**).

Nonaka et al. [19] reported a significantly shorter procedure time in the combination group (dexmedetomidine and propofol) compared to the benzodiazepine group; nevertheless, this finding was lost after pooling with other studies in the analysis, as shown in **Figure 5**.

In terms of safety, our findings support the use of dexmedetomidine as an adjunctive agent in procedural sedation for ESD procedures, consistent with previous studies [20,21]. As noted by Candiotti et al. [20], the Dexmedetomidine group demonstrated a higher incidence of bradycardia, as illustrated in **Figure 7**. However, there was no statistically significant increase in the occurrence of other adverse events such as hypoxia, hypotension, bleeding, or perforation. Additionally, Dexmedetomidine’s cardiovascular and hemodynamic effects are well-known and are attributed to its strong alpha 2-adrenergic agonist effect and include bradycardia, hypotension, and hypoxia [20–23]. Kim et al. evaluated risk factors for dexmedetomidine-associated bradycardia during spinal anesthesia [23] and found that a long tourniquet time and low baseline heart rate were associated with an increased incidence of bradycardia during procedures under spinal anesthesia. Notably, Alshikaria [24] et al. reported that no serious adverse events were observed in patients in the dexmedetomidine group who experienced bradycardia and that their clinical outcomes were not altered due to it, which is also consistent with previous literature [25,26].

The use of Dexmedetomidine as an adjunctive sedative has shown promising results in our meta-analysis, yet this type of intervention needs further exploration. The included studies in this review have already explored the combination of Dexmedetomidine with the two main sedatives, propofol and midazolam. The results are extraordinary in terms of restlessness and bradycardia incidence, the latter being a good sign of less stress and discomfort during the procedure. Less movements (restlessness) during the ESD procedure leads to more convenient and accurate procedures from the operator. So, this therapy should be explored more to reach the best results possible for the patient. More exploration means more multicenter randomized controlled trials and observational studies comparing this type of adjunctive therapy with other adjunctive sedatives and even other types of pain management methods, like local anesthesia in the region of intervention, to test this intervention’s safety and efficacy to standardize its use during ESD procedures in the near future.

Limitations: To our knowledge, this is the first meta-analysis to assess the safety and efficacy of Dexmedetomidine as an adjunctive sedative after ESD. Additionally, we performed subgroup analysis according to each general therapy. Most included articles (6 out of 8) did not conduct a head-to-head comparison between dexmedetomidine and other agents. Also, the limited number of published clinical trials and the number of patients included in certain subgroups make our evidence and conclusions limited on some outcomes. All of our eight included trials were conducted in eastern Asia, including 5 in Japan, two in Korea, and one in China. Thus, the generalizability of this study results to other regions with different ethnicities and medical environments may be affected. A standardized dosage of dexmedetomidine as an adjuvant sedative has not yet been established, resulting in a wide variety of dexmedetomidine regimens.

## 5. Conclusions

In conclusion, our meta-analysis supports the safe use of dexmedetomidine as an adjunctive sedative in ESD procedures. Dexmedetomidine, when combined with other sedatives, appears to reduce restlessness without increasing the risk of hypoxia, hypotension, bleeding, or perforation. The increased risk of bradycardia noted with dexmedetomidine can be perceived as less physiological stress and tachycardia during procedures. However, our findings are limited by the lack of direct comparisons with other sedatives and the predominantly Eastern Asian study populations. Further research, including multicenter trials, is needed to establish optimal dosing regimens and evaluate dexmedetomidine’s efficacy compared to other sedatives and pain management methods in diverse patient populations.

## Data Availability

N/A

## 6. Declarations

### IRB Approval

N/A.

### Ethical Approvals

N/A.

### Consent for Participation

N/A.

