## Appendix 1 for "Dexmedetomidine as an Adjunctive Sedative in Patients Undergoing Endoscopic Submucosal Dissection: A Systematic Review and Meta-Analysis"

| Database | Search Terms | Search Field | Search Results |
| --- | --- | --- | --- |
| Medline | ("endoscopic submucosal dissection" OR "ESD" OR "endoscopic dissection") AND ("dexmedetomidine" OR "Dexmedetomidine Hydrochloride" OR “Precedex” OR "MPV-1440" OR "MPV 1440" OR "MPV1440") | All Field | 17 |
| Cochrane | ("endoscopic submucosal dissection" OR "ESD" OR "endoscopic dissection") AND ("dexmedetomidine" OR "Dexmedetomidine Hydrochloride" OR "MPV-1440" OR "MPV 1440" OR "MPV1440") | All Text | 27 |
| WOS | ((ALL=( ( (endoscopic submucosal dissection OR ESD OR endoscopic dissection)) )) AND  ALL=( ( (Dexmedetomidine OR Dexmedetomidine Hydrochloride OR MPV-1440 OR MPV 1440 OR MPV1440) ) )) | All Fields | 32 |
| SCOPUS | ("endoscopic submucosal dissection" OR "ESD" OR "endoscopic dissection") AND ("dexmedetomidine" OR "Dexmedetomidine Hydrochloride" OR "MPV-1440" OR "MPV 1440" OR "MPV1440") | Title, Abstract, Keywords | 59 |
| EMBASE | Embase: ("endoscopic submucosal dissection" OR "ESD" OR "endoscopic dissection") AND ("dexmedetomidine" OR "Dexmedetomidine Hydrochloride" OR "MPV-1440" OR "MPV 1440" OR "MPV1440") | All Field |  |
