## Appendix 2 for "Dexmedetomidine as an Adjunctive Sedative in Patients Undergoing Endoscopic Submucosal Dissection: A Systematic Review and Meta-Analysis"

| **Author name, year** | **Study design** | **Tool used** | **Overall, ROB** |
| --- | --- | --- | --- |
| **Ashikari, 2021** | RCT | Cochrane RoB 2 | Low |
| **Iwagami, 2023** | Cohort | Newcastle–Ottawa Scale | Low |
| **Kim, 2015** | RCT | Cochrane RoB 2 | Low |
| **Kinugasa, 2018** | RCT | Cochrane RoB 2 | Low |
| **Lee, 2015** | RCT | Cochrane RoB 2 | Low |
| **Luo, 2023** | RCT | Cochrane RoB 2 | Moderate |
| **Nonaka, 2016** | Cohort | Newcastle–Ottawa Scale | Low |
| **Yoshio, 2019** | Cohort | Newcastle–Ottawa Scale | Moderate |

*Table 2 Risk of bias assessment for included studies.*
